# Adverse events following COVID-19 vaccination in young Japanese people: A case-control study of the risk of systemic adverse events by a questionnaire survey

**DOI:** 10.1101/2021.10.01.21264393

**Authors:** Marie Suehiro, Shinya Okubo, Kensuke Nakajima, Kosuke Kanda, Masanobu Hayakawa, Shigeru Oiso, Tsutomu Kabashima, Hideaki Fujita, Yukio Ando, Takahiro Muro

**Affiliations:** Division of Medical Informatics, Department of Pharmacy, Faculty of Pharmaceutical Sciences, Nagasaki International University, Japan; Division of Pharmaceutical Health Care and Science, Department of Pharmacy, Faculty of Pharmaceutical Sciences, Nagasaki International University, Japan; Division of Clinical Pharmaceutics, Department of Pharmacy, Faculty of Pharmaceutical Sciences, Nagasaki International University, Japan; Division of Pharmaceutical Technology, Department of Pharmacy, Faculty of Pharmaceutical Sciences, Nagasaki International University, Japan; Section of Functional Morphology, Department of Pharmacy, Faculty of Pharmaceutical Sciences, Nagasaki International University, Japan; Division of Amyloidosis Research, Department of Pharmacy, Faculty of Pharmaceutical Sciences, Nagasaki International University, Japan

## Abstract

**What is known and objective:** Racial differences in adverse events following COVID-19 vaccines have not been sufficiently studied. Here, we aimed to study the adverse events of Moderna’s intramuscular COVID-19 vaccine in young Japanese people.

**Methods:** A case-control study was conducted using a questionnaire survey. Risk factors were determined using a multivariable logistic regression model. We also compared the occurrence of systemic adverse events in three pairs (minor and adult; male and female; and occurrence and non-occurrence of adverse events after the first dose). Propensity matching was used to balance variables.

**Results:** We analysed 3,369 data points (1,877 after the first dose and 1,492 after the second dose) obtained from a questionnaire survey of 7,965 vaccinated individuals. Comparing the results of the first and second doses, the incidence of local adverse events did not change significantly; however, the incidence of systemic adverse events increased significantly (p < 0.001). Eighty-three percent of the participants complained of local adverse events, and 65% of participants complained of systemic adverse events. Anaphylaxis occurred in one female student (0.03%). Even when an adverse event occurred, most of the symptoms improved within 3 days. Female sex was associated with systemic adverse events after the first and second doses with odds ratios (ORs) (95% confidence interval, CI) of 2.49 (2.03–3.06), and 1.83 (1.28–2.61), respectively. Age (<20 years: minor) was associated with systemic adverse events after the first dose with an OR of 1.80 (1.44–2.24).

The results of the analysis of six cohorts that were created using propensity score matching showed that the incidence of systemic adverse events at the first dose in females was significantly higher than that in males, and that of minors was significantly higher than that of adults.

**What is new and conclusion:** The results of this study clarified, for the first time, the risk factors for several adverse events from the injection of Moderna’s intramuscular COVID-19 vaccine in young Japanese people.

This study suggests that women, minors who experienced adverse events after the first dose, those who experienced adverse events after the first dose, and those who had adverse events after the second dose, should be aware of adverse events.

**What is known and objective:** The COVID-19 crisis has been spreading worldwide, and the number of infected people is increasing due to the emergence of mutant strains. The COVID-19 vaccine is expected to be effective in preventing coronavirus infection and disease aggravation.

In Japan, priority groups, such as healthcare workers and the older adults (aged 65 and over), were the first to be vaccinated. Workplace vaccinations began on 21 June 2021 at universities and businesses; university employees and students have also been vaccinated.

In June 2021, in Japan, Moderna’s intramuscular injection of the COVID-19 vaccine was used for vaccination in places associated with students and corresponding teaching staff. However, as of September 2021, the Comirnaty intramuscular injection vaccine had been used for healthcare workers and elderly people ahead of other COVID-19 vaccines.

Adverse events of the Comirnaty intramuscular injection vaccine in healthcare workers and elderly people and that of Moderna’s intramuscular COVID-19 vaccine in the defence forces, who were prioritised for administration, are currently being studied. However, adverse events associated with the intramuscular injection of Moderna’s COVID-19 vaccine in young people have not yet been well studied in Japan. ^1,2^. In addition, racial differences in adverse events following coronavirus vaccines have not been sufficiently studied.

We have already reported adverse events associated with the first dose of Moderna’s intramuscular injection of its COVID-19 vaccine. In Japan, the intramuscular injection of Moderna’s COVID-19 vaccine will continue to be used in large-scale inoculation venues such as universities and workplaces.

Therefore, to clarify the adverse events following coronavirus vaccination in the young Japanese population, we conducted a questionnaire survey after the second dose for students, faculty, and staff who belong to an educational foundation, Kyushu Bunka Gakuen.

## Methods

### Design

A case-control study with a questionnaire survey was conducted.

### Study population

The study population included people who were vaccinated with Moderna’s intramuscular COVID-19 vaccine.

This vaccination drive included students attending educational facilities run by the educational foundation Kyushu Bunka Gakuen, universities, junior colleges, cooking training facilities, training facilities for dental hygienists, and high schools.

In addition, the vaccination drive included those employed by local companies related to the educational foundation of Kyushu Bunka Gakuen.

### Data collection

Information regarding early adverse events that occurred immediately after vaccination at the site was obtained from the medical records.

To obtain information on the adverse events that occurred after leaving the site, we conducted a questionnaire survey on a website created for this survey using Google Forms. The Japanese Ethics Guidelines for Epidemiological Studies stipulate that the use of existing medical records in observational studies can be used without the consent of individual patients by disclosing information about the purpose and conduct of the study and guaranteeing the opportunity for refusal. An explanation of this study is provided on the website of the Nagasaki International University. Consent to participate in the study was obtained individually.

In addition to assessing adverse events related to vaccination, we obtained the following: type of adverse event, date of occurrence, period of occurrence, and medication/care.

In addition, other attributes (age, sex, allergy history, and history of adverse events to previous medications) were also obtained. Fever was defined as an increase of more than 1 °C from the usual body temperature.

### Statistical analysis

Patient demographics and answers to the questionnaire were summarised with frequencies and percentages for categorical data and median plus range for continuous data.

We compared the patients who experienced adverse events (AE group) with those who did not experience adverse events (No-AE group) using the Wilcoxon ranked-sum test for continuous variables and Fisher’s exact test for dichotomous variables. Differences were considered significant at p < 0.05.

The incidence was calculated for each adverse event, and Fisher’s exact test was performed to compare the first and second doses.

To estimate the risk factors of adverse events, we first evaluated the odds ratios (ORs), 95% confidence intervals (95% CIs), and p-values of each potential risk factor using unadjusted logistic regression models. Second, we determined the risk factors using a multivariable logistic regression model.

Statistical significance was determined if the 95% CIs did not include 1.00 in logistic analyses.

Candidate predictors were selected according to a literature review and clinical expertise. We selected six variables, including patient demographics (age, sex, allergy history, history of adverse events to past medications, history of local adverse events after the first dose of vaccination, and history of systemic adverse events after the first dose vaccination). Age, a continuous variable, was categorised as minor (<20 years) or adult.

Finally, we compared the occurrence of systemic adverse events in three pairs (minors and adults, male and female, and occurrence and non-occurrence of adverse events after the first vaccine dose). Propensity matching was used to balance variables.^3,4^ One-to-one matching without replacement was completed using nearest neighbour matching within a calliper (0.2 of the standard deviation of the logit of the propensity score^5, 6^).

To estimate the propensity score, we fitted a logistic regression model using variables such as age (minor or adult), allergy history, and history of adverse events to past medications. The occurrence of local or systemic adverse events was also used to estimate propensity scores after the second inoculation.

After matching the propensity scores, the statistical balance between the two groups was evaluated. A standardised difference (Std diff) of <0.1 suggests adequate variable balance.^7^ For comparison of the categorical variables, the McNemar test was employed. All statistical analyses were performed using JMP Pro 16 (SAS Institute Inc., Cary, NC, USA).

## Results

### AEs at the vaccination site

In this vaccination program, 7,965 people (3,998 for the first dose and 3,967 for the first and second doses) were vaccinated. Adverse events occurring at the vaccination site (immediately or within 30 minutes after vaccination) are shown in Table 1.

**Table 1.** Adverse events at vaccination site (immediately after vaccination or within 30 minutes after vaccination)

| Adverse events | 1st dose |  |  |  | 2nd dose |  |  |  | Total |  |  |  |
| --- | --- | --- | --- | --- | --- | --- | --- | --- | --- | --- | --- | --- |
|  | Number of people |  | sex |  | Number of people |  | sex |  | Number of people |  | sex |  |
|  | (N = 3,998) | (%) | female | male | (N = 3,967) | (%) | female | male | (N = 7,965) | (%) | female | male |
| Vagal syncope | 5 | 0.13 | 4 | 1 | 2 | 0.05 | 2 |  | 7 | 0.001 | 2 |  |
| Hyperventilation syndrome | 4 | 0.10 | 4 |  | 2 | 0.05 | 2 |  | 6 | 0.001 | 2 |  |
| Feeling sick | 4 | 0.10 | 3 | 1 | 4 | 0.10 | 3 | 1 | 8 | 0.001 | 3 | 1 |
| Suffocation | 3 | 0.08 | 3 |  | 2 | 0.05 | 2 |  | 5 | 0.001 | 2 |  |
| Nausea | 1 | 0.03 | 1 |  | 2 | 0.05 | 2 |  | 3 | 0.000 | 2 |  |
| Anaphylaxis | 1 | 0.03 | 1 |  |  |  |  |  | 1 | 0.000 |  |  |
| Hyperventilation with involuntary movement | 1 | 0.03 | 1 |  |  |  |  |  | 1 | 0.000 |  |  |
| Severe urticaria | 1 | 0.03 | 1 |  |  |  |  |  | 1 | 0.000 |  |  |
| Total | 20 | 0.5 | 18 | 2 | 12 | 0.3 | 11 | 1 | 32 | 0.004 | 11 | 1 |

After the first dose, the proportion of individuals with adverse events at the vaccination site was 0.5%, and anaphylaxis occurred in one female student (0.03%). After the second dose, the proportion of individuals with adverse events at the vaccination site was 0.3%, and none of them had anaphylaxis.

### Demographic characteristics of participants

Table 2 shows the demographic characteristics of participants of the questionnaire survey. After the first dose, we obtained data from 1,993 participants (response rate: 49.8%). A total of 1,877 subjects who agreed to participate in the present study were enrolled in the analysis. Female participants accounted for 66% of the study population, with a median age of 22 years. Twenty-four percent of the study population was younger than 20 years of age.

**Table 2.** Demographic characteristics of participants.

| Characteristic | 1st dose |  | 2nd dose |  | Total |  |
| --- | --- | --- | --- | --- | --- | --- |
|  | Number of people<br>(N = 1,877) | (%) <sup>a)</sup> | Number of people<br>(N = 1,492) | (%) <sup>a)</sup> | Number of people<br>(N = 3,369) | (%) <sup>a)</sup> |
| Female | 1242 | 66 | 992 | 66 | 2234 | 66 |
| Age, median (range), y | 22 | (18-70) | 21 | (18-64) | 22 | (18-70) |
| Age (minor<sup>b)</sup>) | 444 | 24 | 380 | 26 | 824 | 24 |
| Allergy history | 219 | 12 | 151 | 10 | 370 | 11 |
| Adverse events history* | 118 | 6 | 77 | 5 | 195 | 6 |

a) Data are expressed as No.(%) unless otherwise indicated.
b) minor : < 20 years old
\* for past medication except corona virus vaccination

In addition, after the second dose, we obtained data from 1,504 subjects who responded to the questionnaire survey (response rate: 38%). A total of 1,492 subjects who agreed to participate in the present study were enrolled in the analysis. Female participants accounted for 66% of the study population, with a median age of 21 years. Twenty-six percent of the study population was younger than 20 years of age.

### Adverse events after leaving the vaccination site

The incidence of each adverse event is presented in Table 3. Local and systemic adverse events were generally mild. After the first dose, 82% of the participants complained of local adverse events. Injection site pain was the most common local adverse event (71%). Systemic adverse events occurred in 48% of participants. The most common systemic adverse event was myalgia (34%), followed by general fatigue (31%). In contrast, after the second dose, 85% of the participants complained of local adverse events after the second dose. Injection site pain was the most common local adverse event (69%). Systemic adverse events occurred in 88% of participants. The most common adverse event was fever (81%), followed by general fatigue (75%). Comparing the results of the first and second doses, the incidence of all systemic adverse events increased significantly (p < 0.001). The incidence of local adverse events, such as swelling, thermal sensation, redness, and itching, also increased significantly (p < 0.001).

**Table 3.** Incidence of each adverse event.

| Adverse event | 1st dose |  | 2nd dose |  | Comparison of<br>1st and 2nd dose<br>(P-value)<sup>a)</sup> | Total |  |
| --- | --- | --- | --- | --- | --- | --- | --- |
|  | Number of people<br>(N = 1,877) | (%) | Number of people<br>(N = 1,492) | (%) |  | Number of people<br>(N = 3,369) | (%) |
| Local adverse events |  |  |  |  |  |  |  |
| Any | 1,533 | 82 | 1,275 | 85 | 0.003 | 2,808 | 83 |
| Pain | 1,334 | 71 | 1,027 | 69 | 0.161 | 2,361 | 70 |
| Swelling | 757 | 40 | 784 | 53 | <0.001 | 1,541 | 46 |
| Thermal sense | 641 | 34 | 762 | 51 | <0.001 | 1,403 | 42 |
| Redness | 476 | 25 | 636 | 43 | <0.001 | 1,112 | 33 |
| Itching | 370 | 20 | 399 | 27 | <0.001 | 769 | 23 |
| Induration | 312 | 17 | 264 | 18 | 0.434 | 576 | 17 |
| Systemic adverse events |  |  |  |  |  |  |  |
| Any | 894 | 48 | 1,314 | 88 | <0.001 | 2,208 | 66 |
| Myalgia | 632 | 34 | 754 | 51 | <0.001 | 1,386 | 41 |
| Fatigue | 581 | 31 | 1,118 | 75 | <0.001 | 1,699 | 50 |
| Fever | 416 | 22 | 1,216 | 82 | <0.001 | 1,632 | 48 |
| Headache | 386 | 21 | 785 | 53 | <0.001 | 1,171 | 35 |
| Joint pain | 119 | 6 | 498 | 33 | <0.001 | 617 | 18 |
| Chills | 86 | 5 | 656 | 44 | <0.001 | 742 | 22 |

a) Fisher's exact test

Figure 1 shows the time of onset of AEs, Figure 2 shows the duration of AEs, and the type of treatment for AEs is shown in Figure 3. Injection site pain occurred from day 0 to day 1 and continued for 2 days. However, almost all symptoms improved without any treatment. Eleven percent of the participants used pain medication after the second dose. The symptoms improved within a few days for most adverse events. The onset of systemic adverse events, such as fever and malaise, often occurred the day after injection, and these adverse events resolved within a few days in most people. Systemic symptoms in many people improved with the administration of antipyretic analgesics, such as acetaminophen.

**Fig. 1a.**
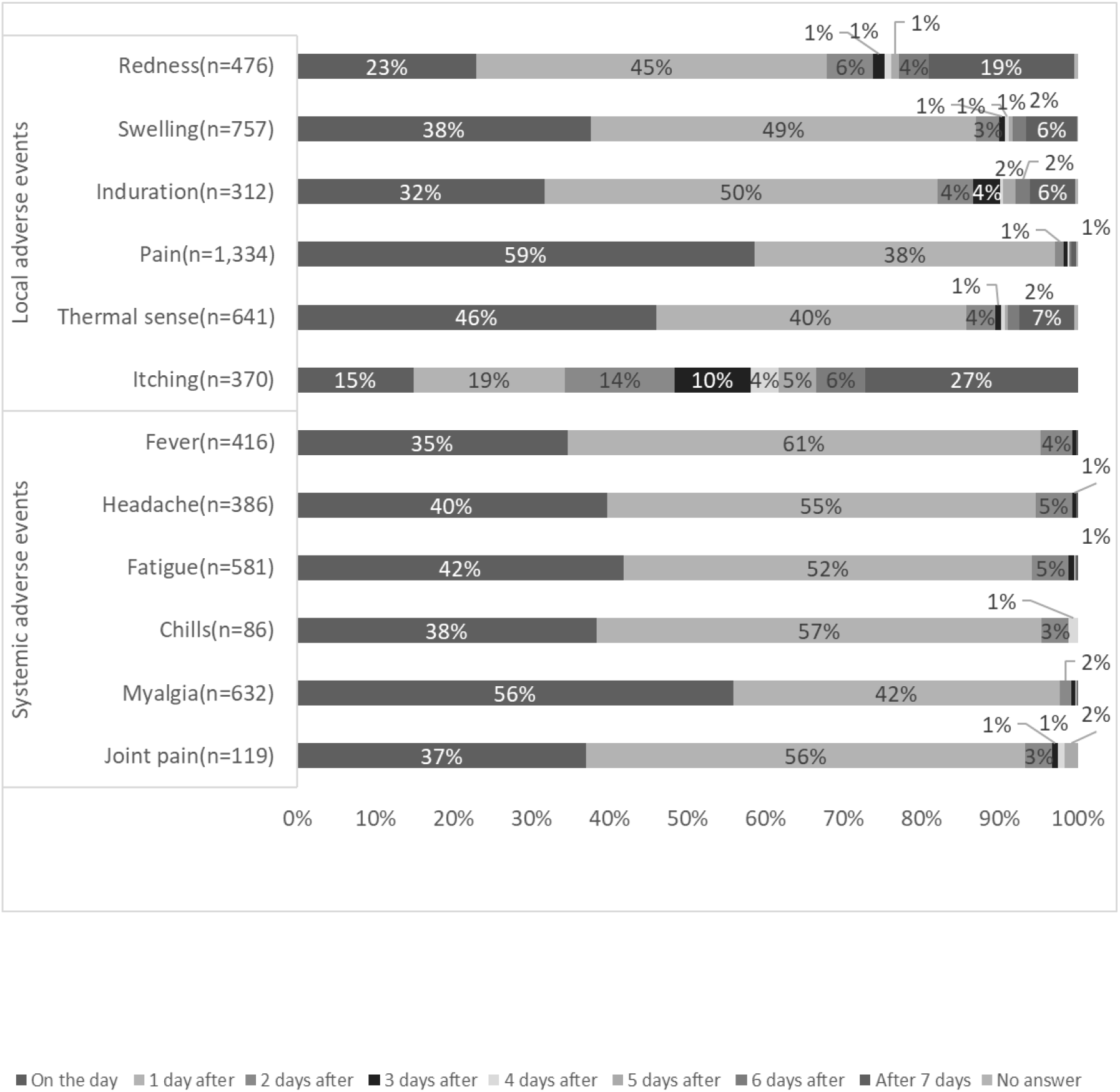
Time of onset of adverse events after the first dose (days after vaccination).

**Fig. 1b.**
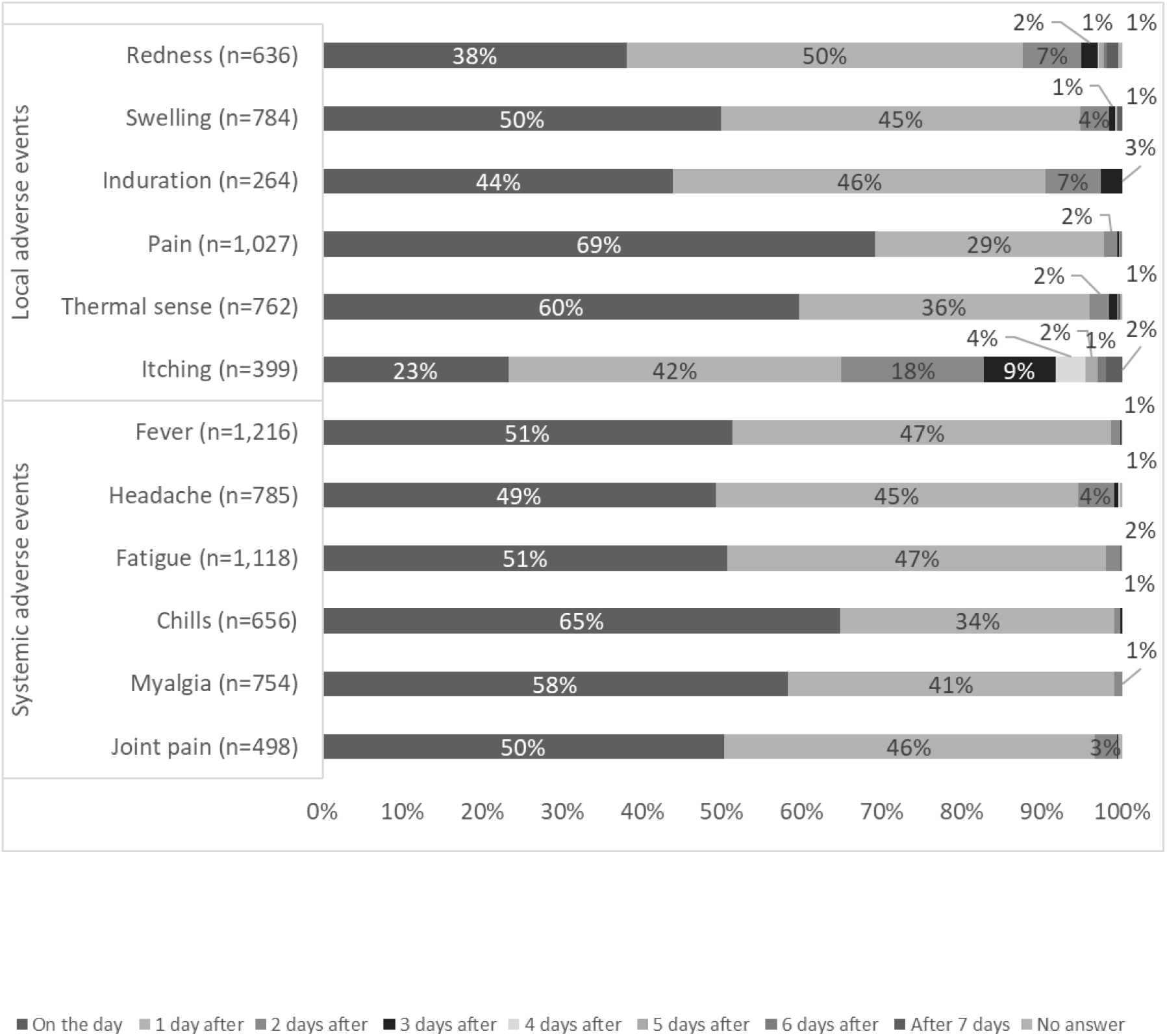
Time of onset of adverse events after the second dose (days after vaccination).

**Fig. 2a.**
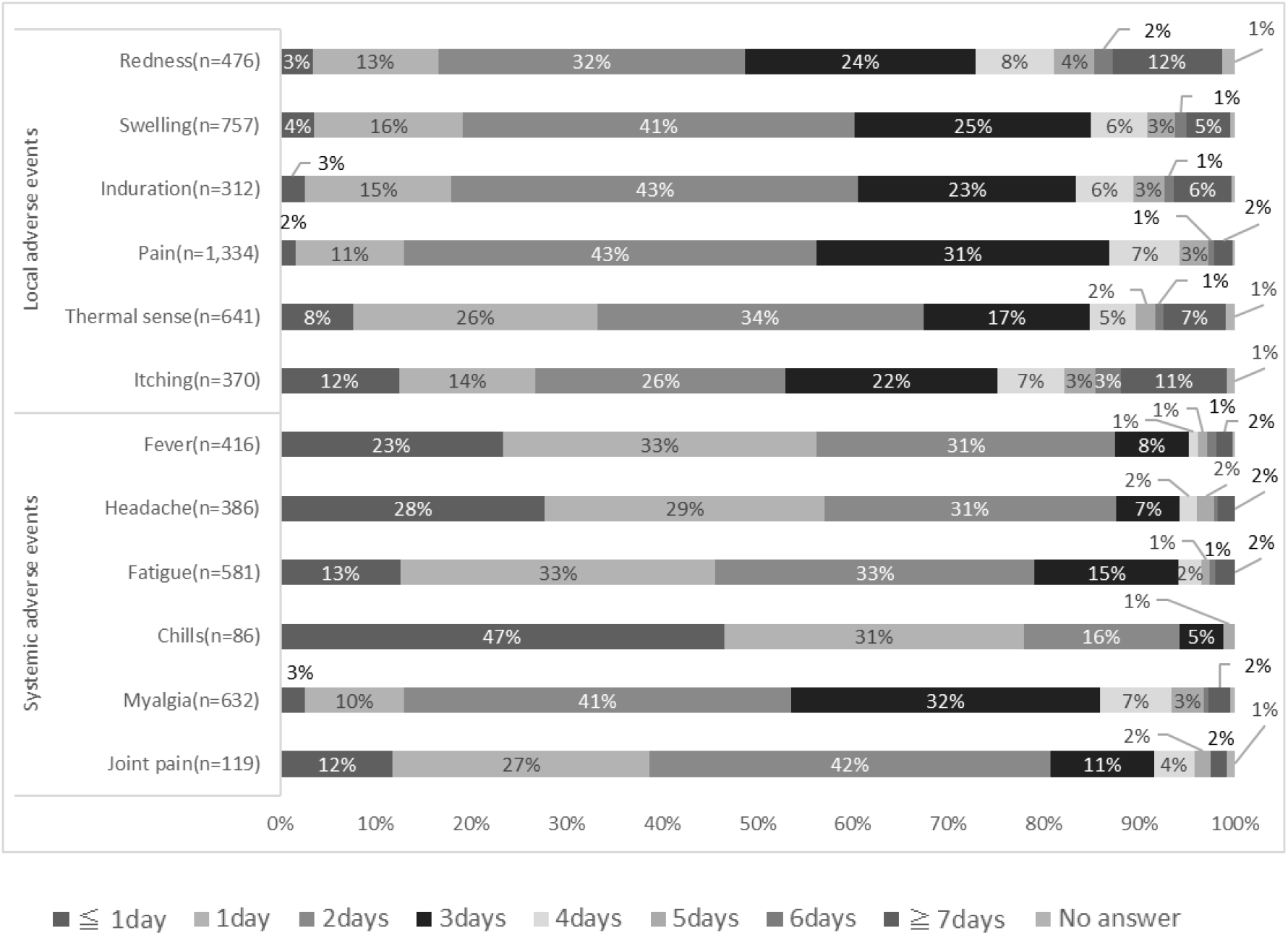
Duration of adverse events after the first dose.

**Fig. 2b.**
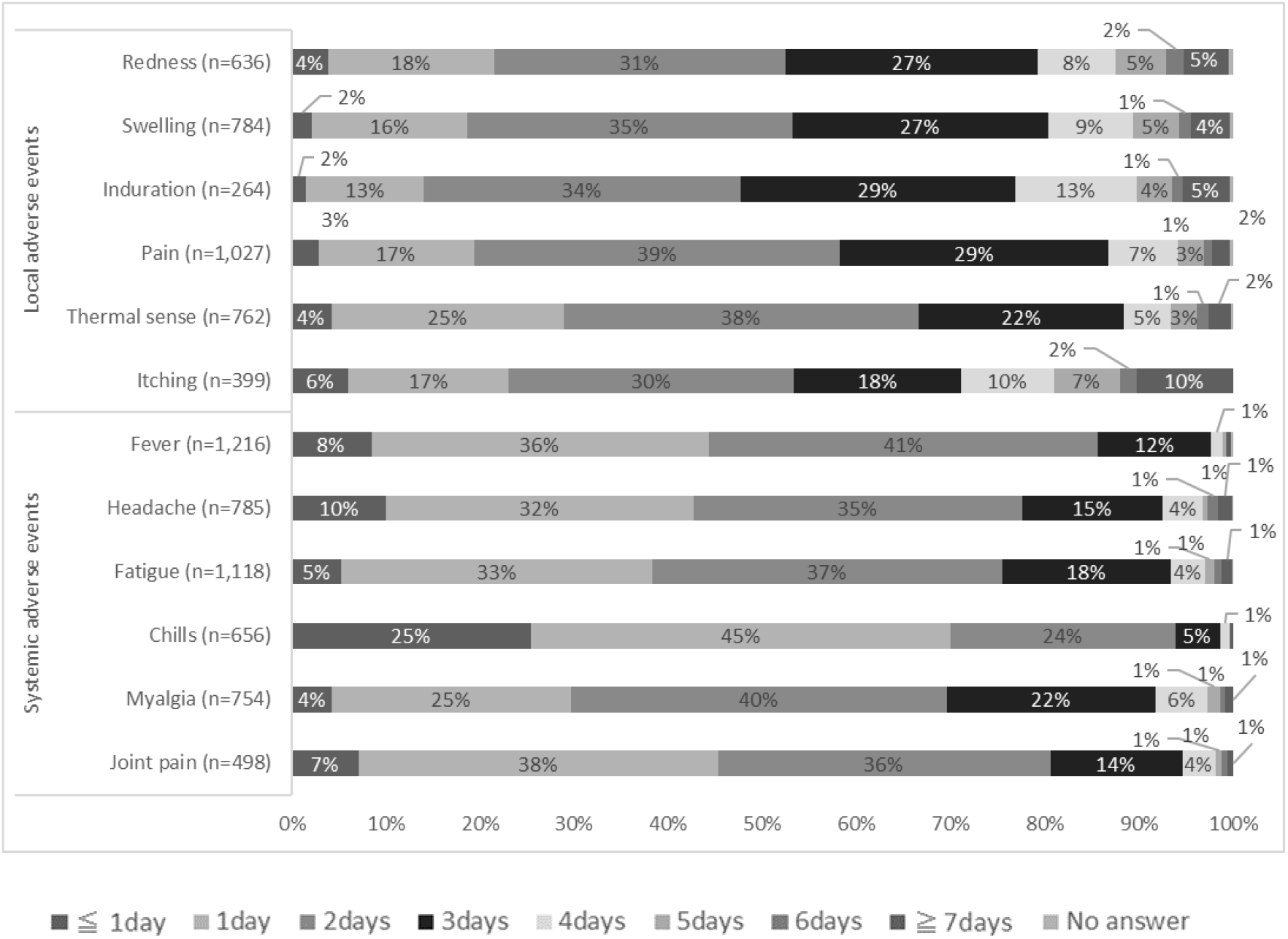
Duration of adverse events after the second dose.

**Fig. 3a.**
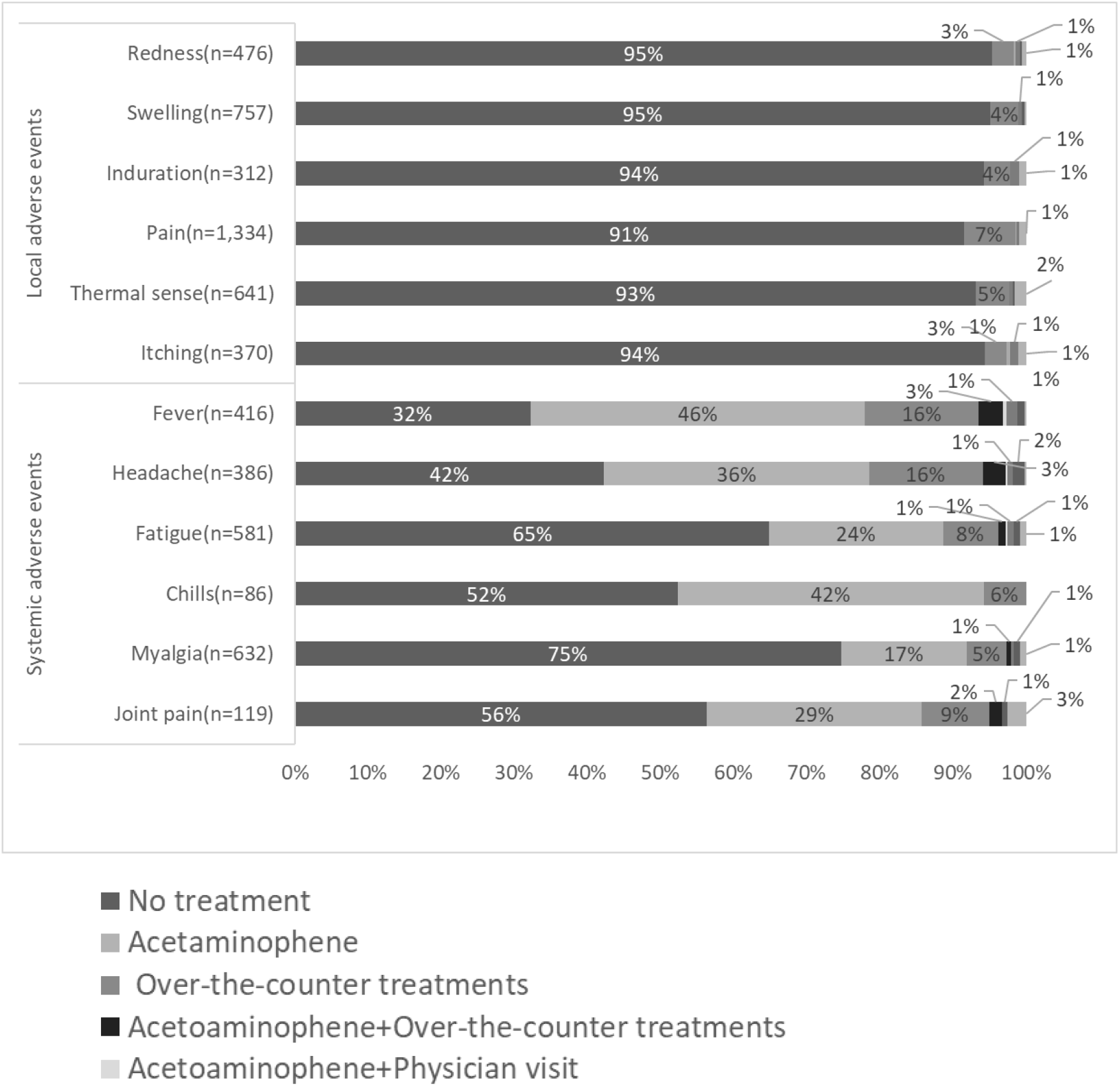
Type of treatment for adverse events after the first dose.

**Fig. 3b.**
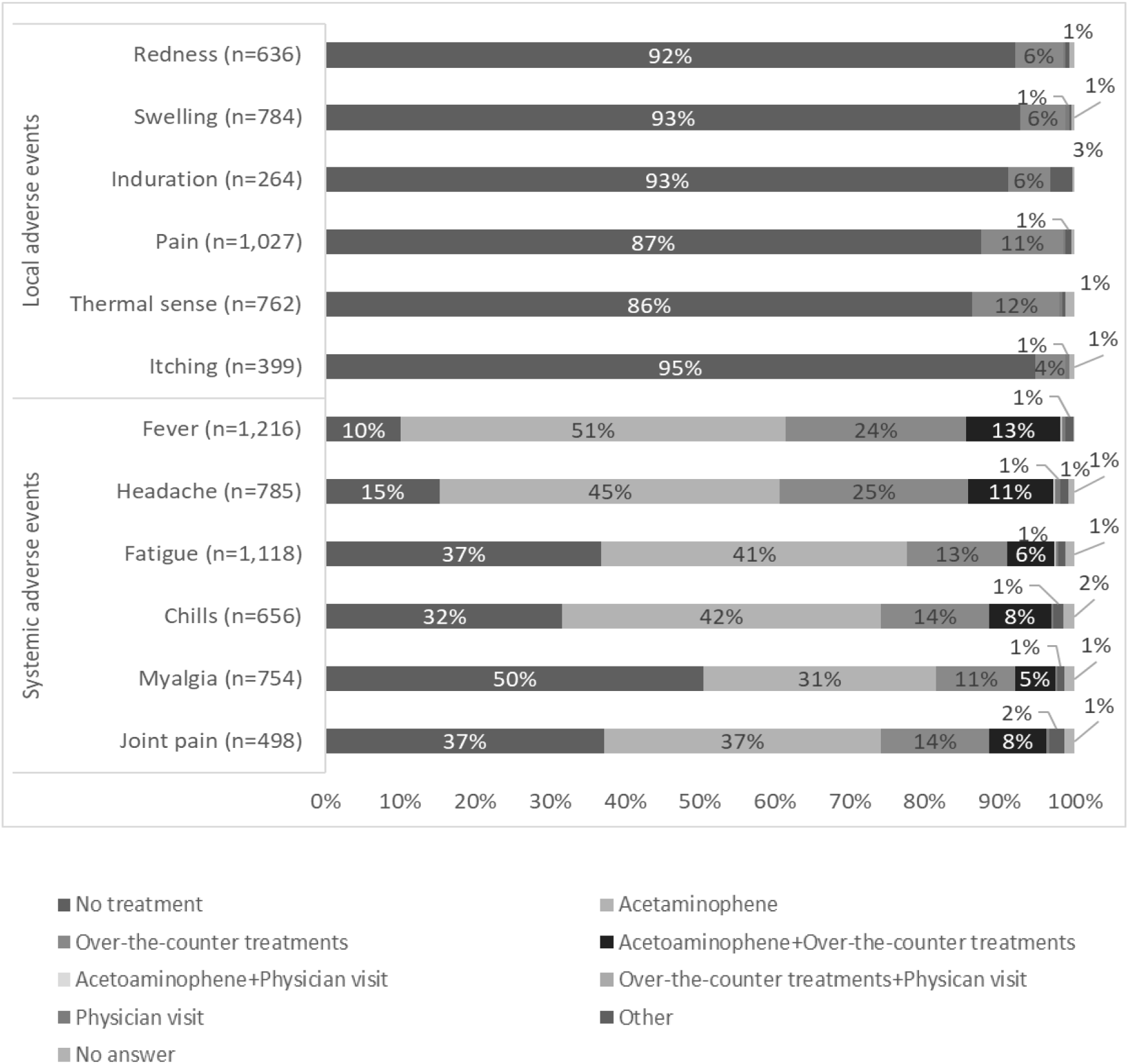
Type of treatment for adverse events after the second dose.

After the first dose, 75% of those who developed symptoms did not undergo any treatment, and 17% were administered acetaminophen. General fatigue often occurred on the day of injection (94%) and continued for 2–3 days thereafter (66%).

In contrast, after the second dose, for those who developed local adverse events, more than 86% did not undergo any treatment, and most showed improvement within 3 days. However, many people who developed systemic adverse events used drugs. In particular, 88% of people who developed fever used antipyretic analgesics, such as acetaminophen. However, most of the symptoms improved within 3 days, and few people visited a medical institution.

### Comparison of the AE and No-AE groups

The characteristics of the AE and no-AE groups are shown in Tables 4 and 5, respectively. Regarding local adverse events, the number of female participants in the AE group was significantly larger than that in the No-AE group (p < 0.001) both after the first and second doses.

**Table 4.** Demographic and clinical characteristics stratified by adverse events on 1st dose^a)^

| **Local adverse events** |  |  |  |
| --- | --- | --- | --- |
| Characteristic | AE group<br>(N = 1,533) | No-AE group<br>(N = 344) | P-Value<sup>b)</sup> |
| sex (female) | 1068 (70%) | 174 (51%) | <0.001 |
| age, median, (range), y | 22 (18-70) | 26 (18-64) | <0.001 |
| age (minor)<sup>c)</sup> | 374 (24%) | 70 (20%) | 0.139 |
| allergy history | 194 (17%) | 25 (7%) | 0.003 |
| adverse events history* | 103 (7%) | 15 (4%) | 0.006 |

| **Systemic adverse events** |  |  |  |
| --- | --- | --- | --- |
| Characteristic | AE group<br>(N = 894) | No-AE group<br>(N = 983) | P-Value<sup>b)</sup> |
| sex (female) | 691 (77%) | 551 (56%) | <0.001 |
| age, median, (range), y | 21 (18-70) | 27 (18-64) | <0.001 |
| age (minor)<sup>c)</sup> | 267 (30%) | 177 (18%) | <0.001 |
| allergy history | 125 (14%) | 94 (10 %) | 0.004 |
| adverse events history * | 71 (8%) | 47 (5%) | 0.111 |

a) Data are expressed as No.(%) unless otherwise indicated.
b) Wilcoxon's rank sum test was performed for continuous variables, and Fisher's exact test was performed for categorical variables.
c) minor : < 20 years old
Abbreviations: AE group, participants who experienced adverse events; No-AE group, participants who did not experience adverse events
\* for past medication except corona virus vaccination

**Table 5.** Demographic and clinical characteristics stratified by adverse events on 2nd dose^a)^

| **Local adverse events** |  |  |  |
| --- | --- | --- | --- |
| Characteristic | AE group<br>(N = 1275) | No-AE group<br>(N = 217) | P-Value<sup>b)</sup> |
| sex (female) | 878 (69%) | 113 (52%) | <0.001 |
| age, median, (range), y | 22 (18-64) | 21 (18-64) | 0.062 |
| age (minor)<sup>d)</sup> | 325 (25%) | 57 (26%) | 0.734 |
| allergy history | 135 (11%) | 16 (7%) | 0.180 |
| adverse events history* | 70 (5%) | 7 (3%) | 0.097 |

| **Systemic adverse events** |  |  |  |
| --- | --- | --- | --- |
| Characteristic | AE group<br>(N = 1314) | No-AE group<br>(N = 178) | P-Value<sup>b)</sup> |
| sex (female) | 909 (69%) | 80 (45%) | <0.001 |
| age, median, (range), y | 22 (18-64) | 30 (18-64) | <0.001 |
| age (minor)<sup>c)</sup> | 342 (26%) | 38 (21%) | 0.349 |
| allergy history | 139 (11%) | 12 (7%) | 0.144 |
| adverse events history* | 73 (6%) | 3 (2%) | 0.027 |

a) Data are expressed as No.(%) unless otherwise indicated.
b) Wilcoxon's rank sum test was performed for continuous variables, and Fisher's exact test was performed for categorical variables.
c) minor: < 20 years old
Abbreviations: AE group, participants who experienced adverse events; No-AE group, participants who did not experience adverse events
\* for past medication except corona virus vaccination

After the first dose, regarding systemic adverse events, the number of female participants and minors in the AE group was significantly higher than that in the No-AE group (p < 0.001) (Table 4).

After the second dose, regarding systemic adverse events, the number of female participants and age in the AE group were significantly higher than those in the No-AE group (p < 0.001) (Table 5).

### Multivariable analysis of AE and No-AE groups

The results of the multivariable analysis of the AE and No-AE groups after the first dose are shown in Table 6.

**Table 6.**
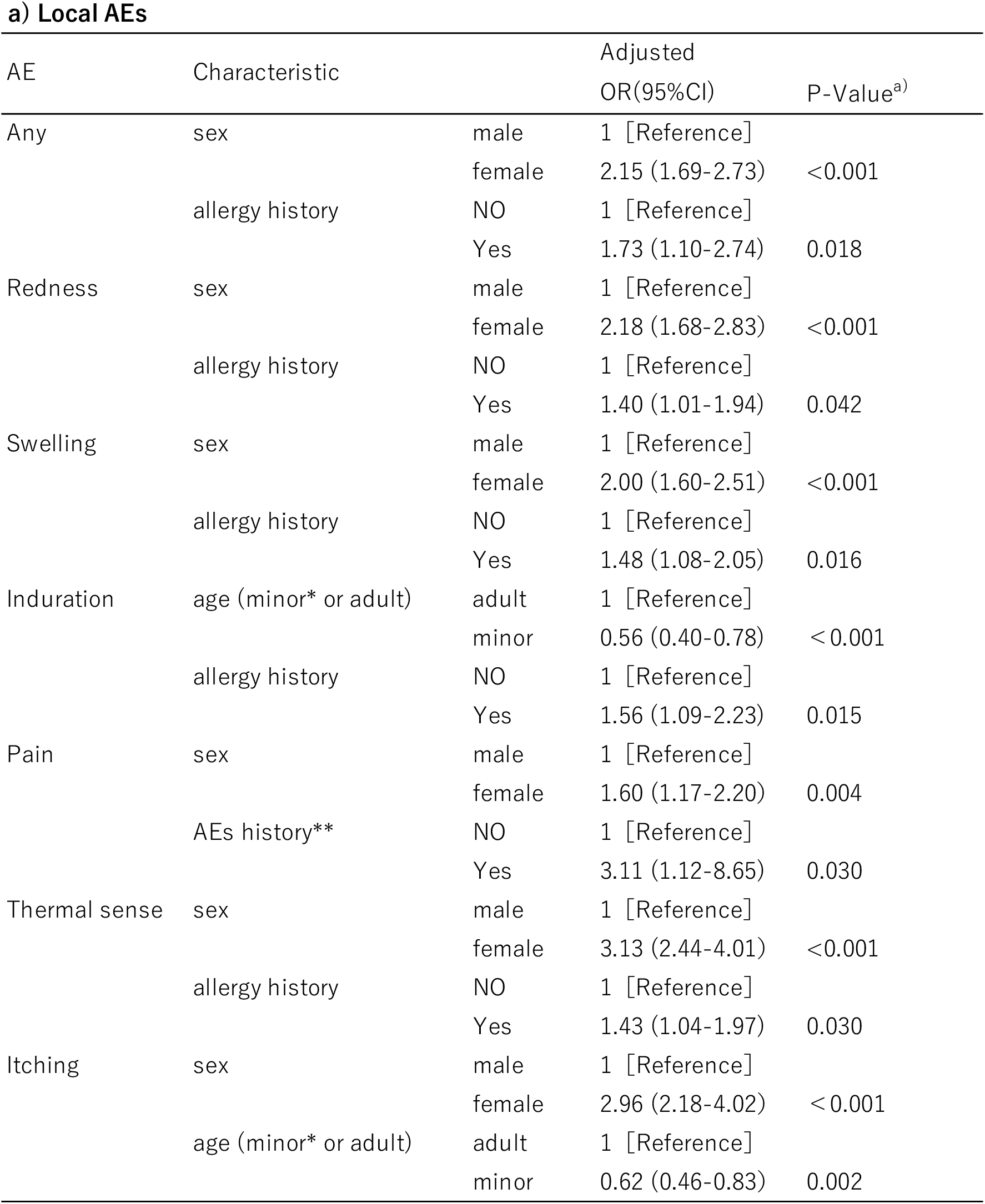
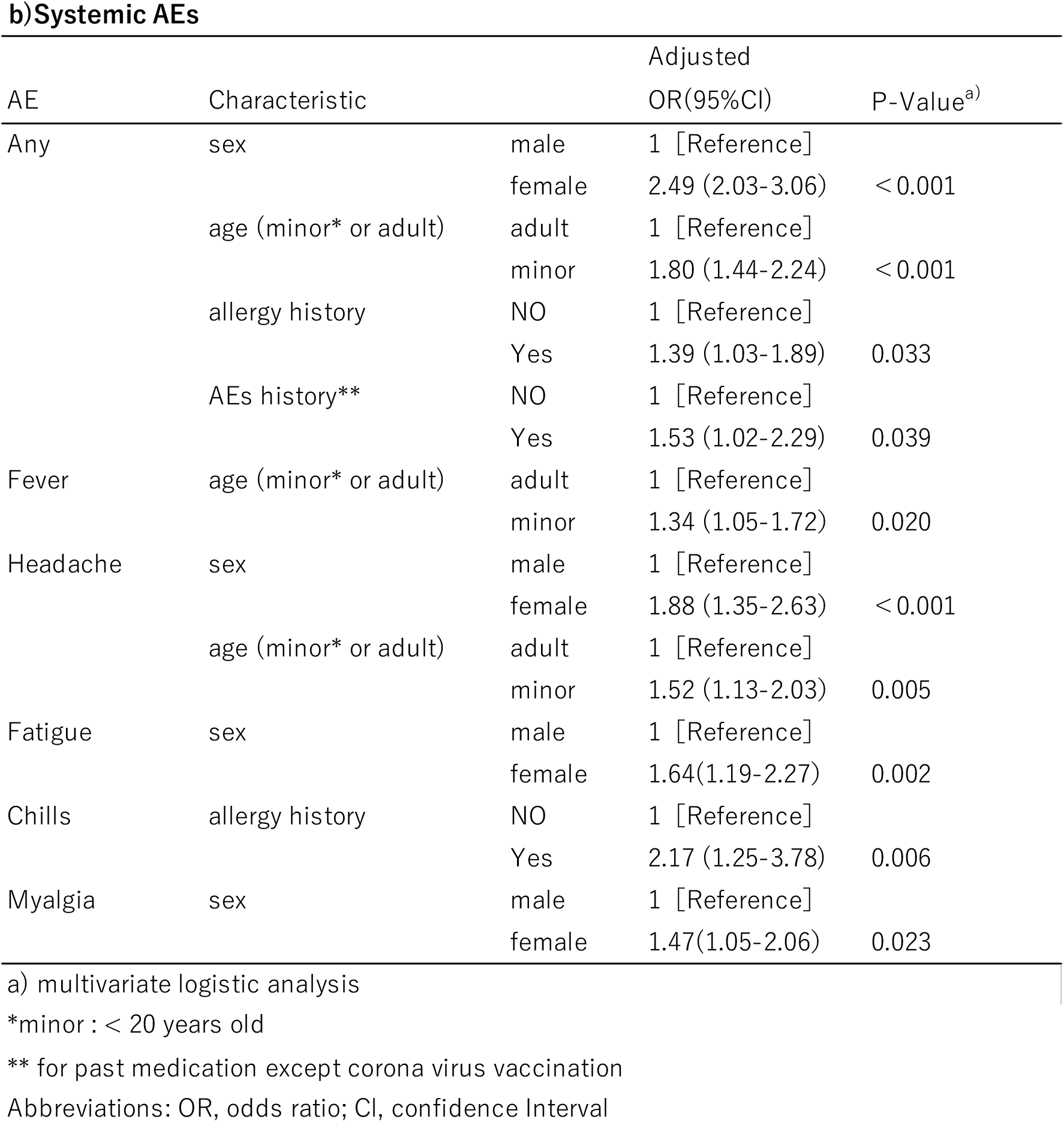
Summary AEs of characteristic by logistic regression analyses on 1st dose.

Local adverse events were associated with sex (female) and allergy history, with ORs (95% CI) of 2.15 (1.69–2.73) and 1.73 (1.10–2.74), respectively (Table 6a). Regarding local adverse events, induration and itching were associated with age (<20 years) (OR [95% CI]: 0.56 [0.40–0.78] and 0.62 [0.46–0.83], respectively), and injection site pain was associated with a history of adverse events with past medication OR (95% CI): 3.11 [1.12–8.65].

Systemic adverse events were associated with sex (female), age (<20 years), allergy history, and history of adverse events with past medications, with ORs (95% CI) of 2.49 (2.03–3.06), 1.80 (1.44–2.24), 1.39 (1.03– 1.89), and 1.53 (1.02–2.29), respectively (Table 6b).

Tables 7(a) and (b) show the results of the multivariable analysis after the second dose. Local adverse events were associated with sex (female) and first dose local adverse event, with ORs (95% CI) of 1.72 (1.60– 2.93) and 18.41 (12.88–26.32), respectively.

**Table 7.**
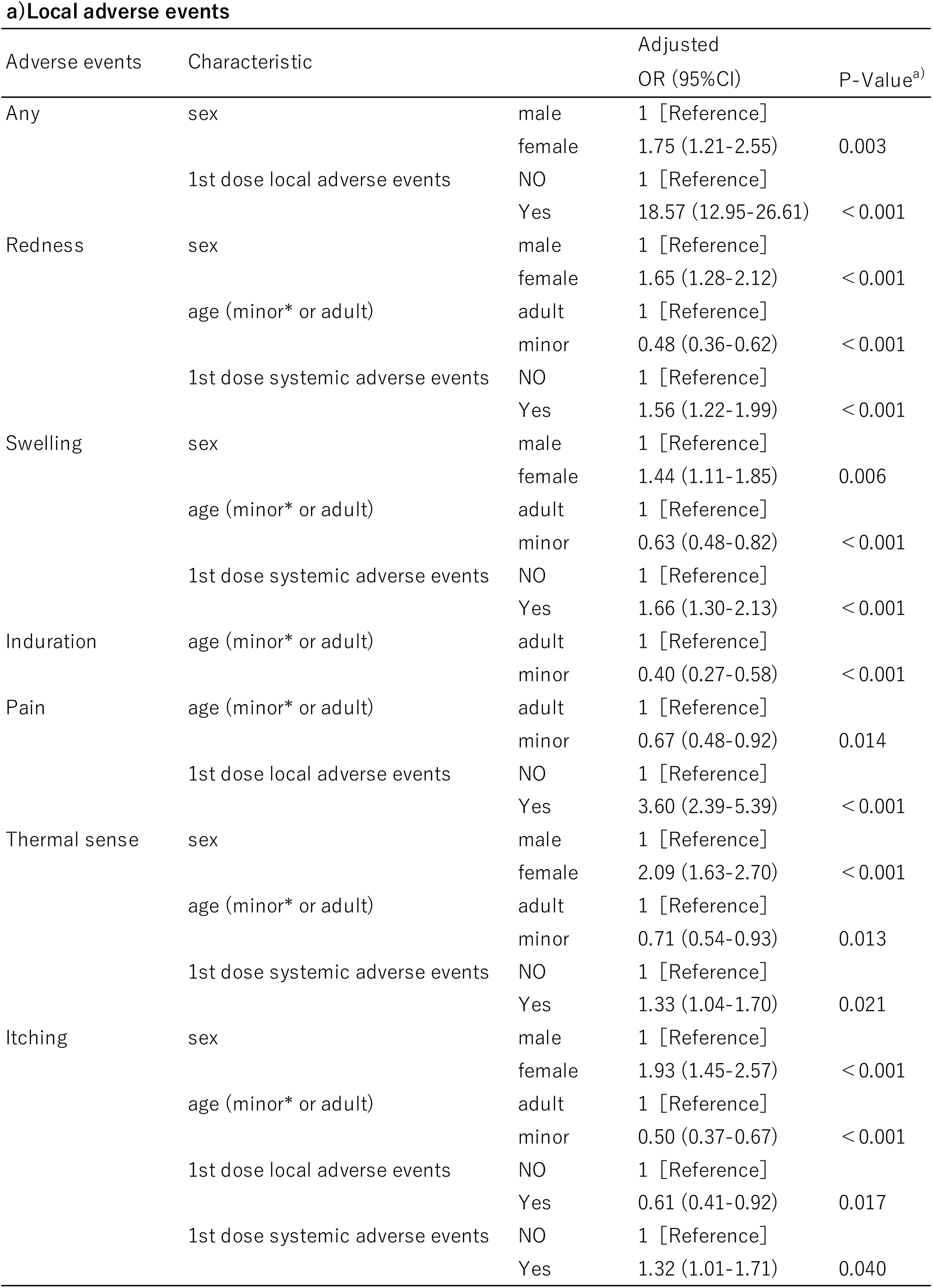
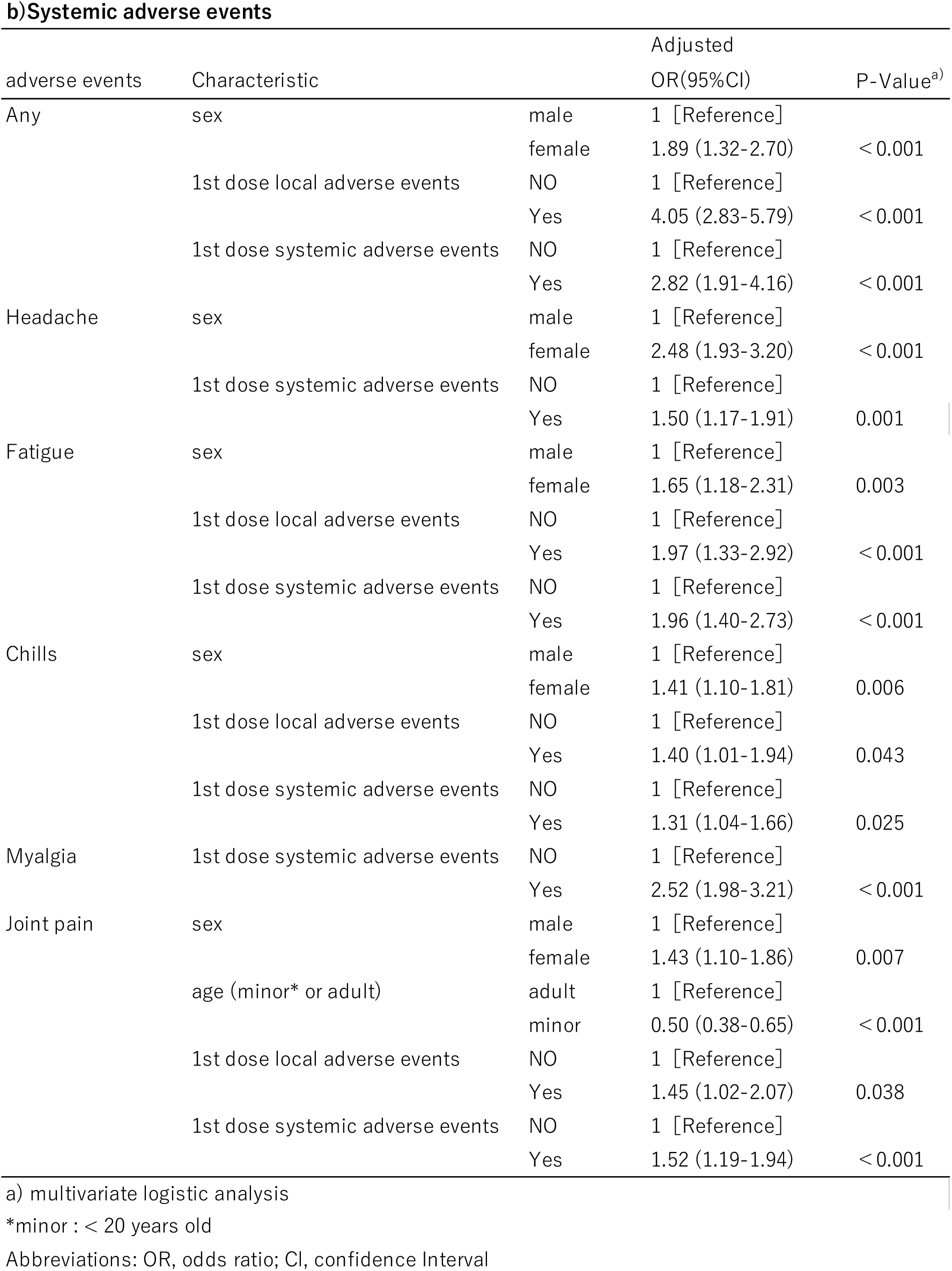
Summary adverse events of characteristic by logistic regression analyses on 2nd dose.

Systemic adverse events were associated with sex (female), first dose local adverse events, and first dose systemic adverse events, with ORs (95% CI) of 1.83 (1.28–2.61), 4.04 (2.83–5.76) and 2.96 (2.01–4.36), respectively.

### McNemar analysis of each risk factor

Table 8 presents the cohorts created for each PSM. Because standardised differences <0.1 were obtained for all variables, the covariate balance in the matched cohort was considerably improved.

**Table 8.** Pre-matching and post-matching summary statistics.

**a) After the 1st dose, sex association for systemic adverse events**
|  | Pre-matching |  |  |  | Post-matching |  |  |  |
| --- | --- | --- | --- | --- | --- | --- | --- | --- |
|  | female<br>(N = 1242) | male<br>(N = 635) | P value<sup>a)</sup> | Std diff | female<br>(N = 628) | male<br>(N = 628) | P value<sup>a)</sup> | Std diff |
| age (minor) | 333 (27%) | 111 (18%) | <0.001 | 0.223 | 111 (18%) | 111 (18%) | 1.000 | 0.000 |
| allergy history | 157 (13%) | 62 (10%) | 0.069 | 0.091 | 60 (10%) | 60 (10%) | 1.000 | 0.000 |
| adverse events history* | 85 (7%) | 33 (5%) | 0.191 | 0.069 | 33 (5%) | 33 (5%) | 1.000 | 0.000 |

**b) After the 1st dose, minor association for systemic adverse events**
|  | Pre-matching |  |  |  | Post-matching |  |  |  |
| --- | --- | --- | --- | --- | --- | --- | --- | --- |
|  | minor<br>(N = 444) | adult<br>(N = 1420) | P value<sup>a)</sup> | Std diff | minor<br>(N = 444) | adult<br>(N = 444) | P value<sup>a)</sup> | Std diff |
| sex (female) | 333 (75%) | 903 (64%) | <0.001 | 0.249 | 333 (75%) | 333 (75%) | 1.000 | 0.000 |
| allergy history | 49 (11%) | 167 (12%) | 0.734 | 0.023 | 49 (11%) | 49 (11%) | 1.000 | 0.000 |
| adverse events history* | 24 (5%) | 94 (7%) | 0.434 | 0.051 | 24 (5%) | 24 (5%) | 1.000 | 0.000 |

**c) After the 2nd dose, sex association for systemic adverse events**
|  | Pre-matching |  |  |  | Post-matching |  |  |  |
| --- | --- | --- | --- | --- | --- | --- | --- | --- |
|  | female<br>(N = 989) | male<br>(N = 503) | P value<sup>a)</sup> | Std diff | female<br>(N = 458) | male<br>(N = 458) | P value<sup>a)</sup> | Std diff |
| age (minor) | 307 (31%) | 75 (15%) | <0.001 | 0.386 | 79 (17%) | 71 (16%) | 0.532 | 0.047 |
| allergy history | 102 (10 %) | 49 (10%) | 0.786 | 0.019 | 44 (10%) | 45 (10%) | 1.000 | 0.007 |
| adverse events history* | 59 (6%) | 18 (4%) | 0.063 | 0.112 | 16 (3%) | 17 (4%) | 1.000 | 0.012 |
| 1st inoculation any local adverse event | 845 (85%) | 379 (75%) | <0.001 | 0.251 | 375 (82%) | 375 (82%) | 1.000 | 0.000 |
| 1st inoculation any systemic adverse event | 646 (65%) | 200 (40%) | <0.001 | 0.529 | 198 (43%) | 197 (43%) | 1.000 | 0.004 |

**d) After the 2nd dose, minor association for systemic adverse events**
|  | Pre-matching |  |  |  | Post-matching |  |  |  |
| --- | --- | --- | --- | --- | --- | --- | --- | --- |
|  | minor<br>(N = 380) | adult<br>(N = 1087) | P value<sup>a)</sup> | Std diff | minor<br>(N = 380) | adult<br>(N = 380) | P value<sup>a)</sup> | Std diff |
| sex (female) | 305 (80%) | 669 (62%) | <0.001 | 0.421 | 304 (80%) | 305 (80%) | 1.000 | 0.000 |
| allergy history | 33 (9%) | 117 (11%) | 0.280 | 0.070 | 33 (9%) | 33 (9%) | 1.000 | 0.012 |
| adverse events history* | 19 (5%) | 54 (5%) | 1.000 | 0.001 | 16 (5%) | 17 (5%) | 1.000 | 0.000 |
| 1st inoculation any local adverse event | 317 (83%) | 890 (82%) | 0.584 | 0.041 | 317 (83%) | 318 (84%) | 1.000 | 0.007 |
| 1st inoculation any systemic adverse event | 264 (69%) | 572 (53%) | <0.001 | 0.351 | 264 (69%) | 264 (69%) | 1.000 | 0.000 |

**e) After the 2nd dose, any local adverse event association for systemic adverse events**
|  | Pre-matching |  |  |  | Post-matching |  |  |  |
| --- | --- | --- | --- | --- | --- | --- | --- | --- |
|  | 1st dose<br>local adverse<br>event<br>(N = 1222) | 1st dose non<br>local adverse<br>event<br>(N = 269) | P value<sup>a)</sup> | Std diff | 1st dose<br>local adverse<br>event<br>(N = 259) | 1st dose non<br>local adverse<br>event<br>(N = 259) | P value<sup>a)</sup> | Std diff |
| sex (female) | 843 (69%) | 146 (54%) | <0.001 | 0.306 | 142 (54%) | 142 (54%) | 1.000 | 0.008 |
| age (minor) | 317 (26%) | 63 (25%) | 0.584 | 0.058 | 63 (24%) | 63 (24%) | 1.000 | 0.009 |
| allergy history | 134 (11%) | 17 (6%) | 0.019 | 0.166 | 17 (7%) | 17 (7%) | 1.000 | 0.000 |
| adverse events history* | 66 (5%) | 10 (4%) | 0.287 | 0.081 | 9 (3%) | 10 (4%) | 1.000 | 0.021 |
| 1st inoculation any systemic adverse event | 758 (62%) | 88 (32%) | <0.001 | 0.614 | 87 (34%) | 87 (34%) | 1.000 | 0.000 |

**f) After the 2nd dose, any systemic adverse event association for systemic adverse events**
|  | Pre-matching |  |  |  | Post-matching |  |  |  |
| --- | --- | --- | --- | --- | --- | --- | --- | --- |
|  | 1st dose<br>systemic<br>adverse<br>event<br>(N = 846) | 1st dose non<br>systemic<br>adverse event<br>(N = 649) | P value<sup>a)</sup> | Std diff | 1st dose<br>systemic<br>adverse<br>event<br>(N = 506) | 1st dose non<br>systemic<br>adverse<br>event<br>(N = 506) | P value<sup>a)</sup> | Std diff |
| sex (female) | 646 (76%) | 343 (53%) | <0.001 | 0.507 | 320 (63%) | 319 (63%) | 1.000 | 0.004 |
| age (minor<sup>b)</sup>) | 264 (32%) | 116 (19%) | <0.001 | 0.314 | 108 (21%) | 107 (21%) | 1.000 | 0.005 |
| allergy history | 99 (12%) | 52 (8%) | 0.024 | 0.124 | 44 (9%) | 45 (9%) | 1.000 | 0.007 |
| adverse events history* | 56 (7%) | 20 (3%) | 0.002 | 0.165 | 16 (3%) | 18 (4%) | 0.862 | 0.022 |
| 1st inoculation any local adverse event | 758 (90%) | 464 (72%) | <0.001 | 0.470 | 438 (87%) | 438 (87%) | 1.000 | 0.000 |

a) Fisher's exact test was performed for categorical variables.
b) minor : < 20 years old
Abbreviations: Std diff, standardized difference
\* for past medication except corona virus vaccination

Table 9 shows a comparison of the incidence of systemic adverse events after the first and second doses.

**Table 9.** Comparisons of systemic adverse events between groups matched by each propensity score.

a) 1st dose
| Group matched by each propensity score | Systemic adverse events |  |
| --- | --- | --- |
|  | N (%)<sup>a)</sup> | P value<sup>b)</sup> |
| Sex |  |  |
| female | 342 (54%) | <0.001 |
| male | 203 (68%) |  |
| Age |  |  |
| minor | 267 (60%) | <0.001 |
| adult | 186 (42%) |  |

a) Data are expressed as No. (%).
b) McNemar test
b) 2nd dose

| Group matched by each propensity score | Systemic adverse events |  |
| --- | --- | --- |
|  | N (%)<sup>a)</sup> | P value<sup>b)</sup> |
| Sex |  |  |
| female | 404 (88%) | 0.022 |
| male | 382 (83%) |  |
| Age |  |  |
| minor | 342 (90%) | 0.701 |
| adult | 345 (91%) |  |
| Local adverse events after 1st dose |  |  |
| Yes | 230 (89%) | <0.001 |
| No | 180 (69%) |  |
| Systemic adverse events after 1st dose |  |  |
| Yes | 474 (94%) | <0.001 |
| No | 432 (85%) |  |

a) Data are expressed as No. (%).
b) McNemar test

After the first dose, female participants and minors had a significantly higher incidence of systemic adverse events which was significantly higher in female participants and minors.

After the second dose, female participants and the occurrence of adverse events after the first dose had a significantly higher incidence of systemic adverse events. However, there were no significant differences between minors and adults.

## What is new and Conclusion

The results of this study clarified, for the first time, the risk factors for several adverse events from the intramuscular injection of Moderna’s COVID-19 vaccine in young Japanese people.

Moderna’s intramuscular COVID-19 vaccine efficacy after the second dose was 94.1% in preventing COVID-19 among those without evidence of previous SARS-CoV-2 infection.^8,9^ Currently, this vaccine requires two doses to be fully effective, with the second dose having a higher frequency of adverse events.^1,2,8-10^ In this study, which compared the occurrence of adverse events after the first and second doses, the results showed that the second dose had an increase in all systemic adverse events and in many local adverse events. This is consistent with the results of other reports.^8,9^

In a previous study, we reported that minors aged <20 years were at greater risk of systemic adverse events from Moderna’s intramuscular injection of the COVID-19 vaccine after the first dose.^11^ However, the same results were not observed for the second dose.

According to a report by the Japanese Defence Forces, local pain incidence tends to be higher in older people.^1^However, other adverse events decrease with increasing age. In comparison with the Comirnaty intramuscular injection, there is less tendency for the incidence to decrease due to older age^1^.

Based on the results of our multivariable logistic analysis, being a minor was found to be an independent influencing factor for systemic adverse events as well as fever after the first dose of Moderna’s intramuscular injection COVID-19 vaccine, but minors were not shown to be independent influencing factors for systemic adverse events and fever. In contrast, adults were more likely to have local adverse events, such as redness, induration, and itching, after the second dose.

Second, the results of this study show that female sex is a major risk factor for many adverse events, except for local pain and induration.

Previous reports showed that women are more likely to develop anaphylaxis immediately after vaccination than men.^12^ Similarly, in the present study, more women than men had symptoms immediately after vaccination. The reasons why female participants have stronger immunity and a higher incidence of autoimmune diseases are not clear. However, sex hormones, such as oestrogen, contribute to the development and activity of the immune system, accounting for differences in gender-related immune responses.^13^ The ratio of naive B cells before vaccination and the ratio of activated CD8-positive T cells after vaccination are detected as immunological features that positively correlate with the increase in antibody titre after vaccination.^14^ The results of this study showed that female participants had a higher incidence of adverse events.

According to a study by the Chiba University in Japan, the older the age, the lower the antibody titre, and it has been reported that women of all ages have higher antibody titres.^15^ Furthermore, from the results of the multivariable analysis, it was reported that the female sex is a factor that tends to increase antibody titres after vaccination, and that older age is also a factor because antibody titres do not easily increase.^14^ The relationship between antibody titres and side reactions was not investigated in the present study. This may need to be analysed in detail in the future.

In this study, to obtain information on AEs that occurred after leaving the vaccination site, we conducted a questionnaire survey on a website. Therefore, the existence of a reporting bias cannot be completely ruled out. This is one of the limitations of the present study.

This study suggested that women, minors who experienced adverse events after the first dose, those who experienced adverse events after the first dose, and those who had adverse events after the second dose, should be aware of adverse events.

However, even if an adverse event occurs, most of the symptoms improve within 3 days, and the adverse events can be appropriately managed by taking antipyretic analgesics such as acetaminophen. The COVID-19 vaccine has also been shown to be effective against mutant strains, such as the delta strain.^16^ COVID-19 has a higher risk of severe effects than post-vaccination adverse events, and vaccination is now recommended for patients with heart disease and pregnant women.^17,18^ The population does not need to be overly afraid of adverse events, and it is recommended that those at high risk make as many advanced preparations as possible, such as having antipyretic analgesics at hand, food for several days, and taking a few days off after vaccination. It is important to note that people must be vaccinated.

## Data Availability

Due to the nature of this research, participants of this study did not agree for their data to be shared publicly, so supporting data is not available.

## References

1) Japan Ministry of Health, Labour and Welfare. Website https://www.mhlw.go.jp/stf/seisakunitsuite/bunya/vaccine_kenkoujoukyoutyousa.html (Accessed on: 2021/08/17)

2) Izumo T, Kuse N, Awano N, et al. Side effects and antibody titer transition of the BNT162b2 messenger ribonucleic acid coronavirus disease 2019 vaccine in Japan. Respir Investig 2021;59:635–642

3) Imbens GW, Rubin DB. Causal inference in statistics, social, and biomedical sciences. New York: Cambridge University Press; 2015. 592 p.

4) Austin PC, Schuster T. The performance of different propensity score methods for estimating absolute effects of treatments on survival outcomes: A simulation study. Stat Methods Med Res 2014;25:2214–2237.

5) Austin PC. Optimal caliper widths for propensity-score matching when estimating differences in means and differences in proportions in observational studies. Pharm Stat 2011;10:150–161.

6) Austin PC. A comparison of 12 algorithms for matching on the propensity score. Stat Med 2014;33:1057–1069.

7) Normand ST, Landrum MB, Guadagnoli E, et al. Validating recommendations for coronary angiography following acute myocardial infarction in the elderly: a matched analysis using propensity scores. J Clin Epidemiol 2001;54:387–398.

8) Baden LR, El Sahly HM, Essink B, et al. Efficacy and safety of the mRNA-1273 SARS-CoV-2 vaccine. N Engl J Med 2021;384:403–416.

9) Chapin-Bardales J, Gee J, Myers T. Reactogenicity following receipt of mRNA-based COVID-19 vaccines. JAMA 2021;325:2201–2202.

10) Oliver SE, Gargano JW, Marin M, et al. The Advisory Committee on Immunization Practices’ Interim Recommendation for Use of Moderna COVID-19 Vaccine — United States, December 2020. MMWR Morb Mortal Wkly Rep 2021;69:1653–1656.

11) Suehiro M, Okubo S, Nakajima K, et al. Adverse events following COVID-19 virus vaccination in Japanese young population: The first cross-sectional study conducted by a questionnaire survey after the first-time-injection. medRxiv 2021.07.23.21261029; DOI: 10.1101/2021.07.23.2126102

12) Shimabukuro T. Allergic reactions including anaphylaxis after receipt of the first dose of Moderna COVID-19 vaccine — United States, December 21, 2020–January 10, 2021. Am J Transplant 2021;21:1326–1331.

13) Moulton VR. Sex hormones in acquired immunity and autoimmune disease. Front Immunol 2018;9:2279.

14) Kageyama T, Tanaka S, Etor K, et al. Immunological features that determine the strength of antibody responses to BNT162b2 mRNA vaccine against SARS-CoV-2. Preprint at bioRxiv https://doi.org/10.1101/2021.06.21.449182 (2021)

15) Kageyama T, Ikeda K, Tanaka S, et al. Antibody responses to BNT162b2 mRNA COVID-19 vaccine in 2,015 healthcare workers in a single tertiary referral hospital in Japan. Preprint at medRxiv https://doi.org/10.1101/2021.06.01.21258188 (2021)

16) Bernal JL, Andrews N, Gower C, et al. Effectiveness of Covid-19 vaccines against the B. 1.617. 2 (Delta) variant. N Engl J Med 2021;385:585–594.

17) Barda N, Dagan N, Ben-Shlomo Y, et al. Safety of the BNT162b2 mRNA Covid-19 vaccine in a nationwide setting. N Engl J Med 2021;385:1078–1090.

18) Shimabukuro TT, Kim SY, Myers TR, et al. Preliminary findings of mRNA Covid-19 vaccine safety in pregnant persons. N Engl J Med 2021; 384:2273–2282

